# Puberty timing and sex-specific trajectories of systolic blood pressure: a prospective cohort study

**DOI:** 10.1101/2021.01.14.21249799

**Authors:** Kate N O’Neill, Joshua A Bell, George Davey Smith, Kate Tilling, Patricia M Kearney, Linda M O’Keeffe

## Abstract

**Background:** Sex differences in systolic blood pressure (SBP) emerge during adolescence but the role of puberty is not well understood. We examined sex-specific changes in SBP preceding and following puberty and examined the impact of puberty timing on SBP trajectories in females and males.

**Methods:** Trajectories of SBP before and after puberty and by timing of puberty in females and males in a contemporary English birth cohort study were analysed. Repeated measures of height from age 5 to 20 years were used to identify puberty timing (age at peak height velocity). SBP was measured on ten occasions from 3 to 24 years (N participants=4,062, repeated SBP measures=29,172). Analyses were preformed using linear spline multilevel models based on time before and after puberty and were adjusted for parental factors and early childhood factors including BMI gain from age one up to 9 years.

**Results:** Mean age at peak height velocity was 11.7 years (standard deviation (SD) =0.8) for females and 13.6 years (SD=0.9) for males. In adjusted models, females and males had similar SBP at age 3. Males had faster rates of increase in SBP before puberty leading to 10.19mmHg (95% CI: 6.80, 13.57) higher mean SBP at puberty which remained similar at 24 years [mean difference; 11.43mmHg (95% CI: 7.22, 15.63)]. Puberty timing was associated with small transient differences in SBP trajectories post-puberty in both sexes and small differences at 24 years in females only.

**Conclusion:** A large proportion of the higher SBP observed in males compared to females in early adulthood is accrued before puberty. Prevention of high SBP in adult males may therefore benefit from a life course approach starting from before puberty. Interventions targeting puberty timing are unlikely to greatly influence SBP in females and males in early adulthood.

## Introduction

High systolic blood pressure (SBP) is a leading modifiable risk factor for cardiovascular disease (CVD) (1–3). SBP tracks from childhood through to adulthood (4); both higher levels of SBP and faster rates of increase in SBP during adolescence are positively associated with the risk of developing hypertension in later life (5). Sex differences in SBP across the life course are well established with males having higher SBP than females throughout much of adult life until mid to later life when steeper rises in SBP are observed in females (6–9). Sex differences in SBP emerge during adolescence and by age 18 there is evidence of 10mmHg higher SBP in males compared to females (6,10–12).

Puberty has been identified as a crucial period in adolescence which may account for the emergence of a sex difference in SBP with the disparate action of sex steroids on blood pressure put forward as a biological mechanism (11,13,14). However, few studies to date have examined and compared change in SBP before and after puberty in females and males (14,15). In an analysis with repeated measures of SBP over a ten-year period from before to after puberty, males had higher SBP than females and similar patterns of change throughout the time observed, albeit with faster rates of increase in SBP in males around the pubertal growth period (15). However, the study included only 182 participants, not all of whom were followed up into early adulthood, limiting insights into the role of puberty in SBP change after more transient effects on SBP at puberty subside.

In addition to the potential role of puberty in the emergence of sex differences in SBP, several studies have examined whether puberty timing influences SBP later in adulthood (16–20). However, results have been largely inconsistent with some studies demonstrating associations between early puberty and higher SBP in both sexes (16,17) while others document associations in males but not females (18,19) or provide no strong evidence of associations in either sex (20). These studies have been limited by their use of self-report measures of puberty timing or have lacked data on pre-pubertal adiposity gain, an important confounder of puberty timing - cardiovascular risk associations (20,21). In addition, these studies only examined single measurements of SBP in adulthood. Understanding whether puberty timing is associated with SBP trajectories before and after puberty may provide further insights into the potential causality of associations between puberty timing and SBP in adulthood in females and males. Any observed associations of puberty timing with SBP before puberty are unlikely to reflect a causal effect as this is temporally implausible and therefore likely explained by confounding and/or possibly shared genetic architecture between SBP and puberty timing. Consequently, if puberty timing is associated with SBP measured before puberty to a similar degree as SBP measured in adulthood (after puberty), this would suggest that puberty timing itself is unlikely to be a cause of SBP.

Using an objective growth-based measure of puberty (age at peak height velocity, aPHV), repeated SBP measures from 3-24 years of age from a large contemporary prospective birth cohort study in the southwest of England and with adjustment for pre-pubertal adiposity gain, we first examine change in SBP before and after puberty to better understand whether sex-specific changes in SBP precede or follow puberty. Second, we examine the association between puberty timing and SBP trajectories before and after puberty in females and males, to gain a better understanding of the likely causality of associations between puberty timing and SBP in adulthood.

## Methods

### Study participants

Data were from first-generation offspring of the Avon Longitudinal Study of Parents and Children (ALSPAC), a population-based prospective birth cohort study in southwest England (22,23). Pregnant women resident in one of the three Bristol-based health districts with an expected delivery date between April 1, 1991 and December 31, 1992 were invited to participate. The study is described elsewhere in detail (22–24). ALSPAC initially enrolled a cohort of 14,451 pregnancies, from which 14,062 live births occurred and 13,988 children were alive at 1 year of age. When the oldest children were approximately 7 years of age, an attempt was made to bolster the initial sample with eligible cases who had not joined the study originally. Therefore, the total sample size for analyses using any data collected after the age of 7 is 15,454 pregnancies, resulting in 15,589 foetuses. Of these 14,901 were alive at 1 year of age. Follow-up has included parent- and child-completed questionnaires, research clinic attendance, and links to routine data. Data gathered from participants at 22 years of age and onwards were collected and managed using REDCap (Research Electronic Data Capture) electronic data capture tools (25,26). Ethical approval for the study was obtained from the ALSPAC Ethics and Law Committee and the Local Research Ethics Committees. Informed consent for the use of data collected via questionnaires and clinics was obtained from participants following the recommendations of the ALSPAC Ethics and Law Committee at the time. The study website contains details of all the data that is available through a fully searchable data dictionary http://www.bristol.ac.uk/alspac/researchers/our-data/.

### Data

#### Assessment of puberty timing

Puberty is a period of intense hormonal activity and rapid growth, of which the most striking feature is the spurt in height (27). Age at peak height velocity (aPHV) is a validated measure of pubertal timing (27) captured using Superimposition by Translation and Rotation (SITAR), a non-linear multilevel model with natural cubic splines which estimates the population average growth curve and departures from it as random effects (28,29). Using SITAR, PHV was identified in ALSPAC participants using numerical differentiation of the individually predicted growth curves, with aPHV being the age at which the maximum velocity is observed (28–30). Repeated data on measured height from research clinics were used here to derive aPHV. Individuals with at least one measurement of height from 5 to <10 years, 10 to <15 years and 15 to 20 years are included here. Data were analysed for females and males separately. The model was fitted using the SITAR package in R version 3.4.1. Further details of height measures are included in Supplementary eTable 1 and information on how aPHV was derived is described elsewhere (30) and in Supplementary Material eMethods 1.

#### Measurement of systolic blood pressure

Ten measurements of SBP (mean ages 3, 5, 7, 9, 10, 11, 12, 15, 18 and 24 years) were available from research clinic assessments. In a random 10% of the cohort, SBP was measured at ‘Children in Focus’ clinical assessments conducted in early childhood (ages 3, 4, 5 years) (22). After this (from 7 to 24 years), all children were invited to attend focus clinics. At each clinic, SBP was measured at least twice each with the child sitting and at rest with the arm supported, using a cuff size appropriate for the child’s upper arm circumference and a validated blood pressure monitor. The mean of the two final measures was used. Further details are provided in the Supplementary Material eMethods 2.

#### Measurement of covariates

We considered the following as potential confounders of the association between puberty/age at puberty and SBP: birth weight, gestational age, maternal education, mother’s partner’s education, parity, maternal smoking during pregnancy, maternal age, maternal pre-pregnancy body mass index (BMI), household social class, marital status, ever breastfed (all measured by mother- or mother’s partner-completed questionnaires) and pre-pubertal gains in BMI from one up to nine years of age. Further details of measurements are available in Supplementary Material eMethods 3.

### Sample size for analysis

Participants who had a measure of aPHV, at least one measure of SBP from 3 to 24 years and complete data on all confounders were included in analyses, leading to a total sample of 4,062 (2,139 females and 1,923 males). Participants who reported being pregnant at the 18-year clinic or 24-year clinic were excluded from the multilevel models at that time point only (N=9).

### Statistical analysis

#### Pubertal age-based multilevel model

Linear spline multilevel models were used to examine change in SBP during childhood and adolescence (31,32). Using terms such as splines to account for non-linearity in the trajectory, such models can estimate mean trajectories of the outcome while accounting for the non-independence or clustering of repeated measurements within individuals, change in scale and variance of measures over time, and differences in the number and timing of measurements between individuals (using all available data from all eligible participants under a missing at-random assumption (MAR)) (33,34). A common approach to modelling change over time using multilevel models involves examining change by chronological age (10,35). However, when change before or after a specified event is of interest (for example, puberty or menopause), it is also possible to model change according to other time metrics such as time before and/or after the event (21). Thus, to gain a greater understanding of the role of puberty and its timing in change in SBP during childhood and adolescence, we modelled trajectories of SBP by time before and after puberty. The final model for females had four periods of SBP change: one pre-pubertal period and three post-pubertal periods. In males, the final model for change in SBP also had four periods of change: two pre-pubertal periods and two post-pubertal periods. Due to different periods of change in females and males all models were sex stratified. Further details on the selection of models and model fit are included in Supplementary Material eMethods 3, eTable 2 and eTable 3.

To explore sex-specific change in SBP before and after puberty, we compared SBP trajectories for the median female (aPHV = 11.6 years) and male (aPHV = 13.6 years); this provided insight into the sex-specific changes in SBP preceding and following puberty in females and males. As a female with the median aPHV is younger chronologically than the median male and SBP increases with age, we also compared SBP trajectories for a female and male with similar aPHV (age 12.8 years in females [90^th^ percentile] and 12.4 years in males [10^th^ percentile]). This provided insights into whether any differences in trajectories, particularly differences in SBP at puberty between the median female and male were independent of differences in chronological age. We compared the difference in SBP between females and males at age 3, at puberty and age 24 by calculating the mean difference between the sexes and using the pooled standard error to calculate 95% confidence intervals for the difference.

We then examined the effect of aPHV on SBP trajectories before and after puberty in females and males separately. Differences in the rate of change in SBP before and after puberty by aPHV were explored by including an interaction between centred sex-specific aPHV and the intercept (SBP at puberty) and each linear spline period. Figures presented compare SBP trajectories for the median, 10^th^ and 90^th^ sex-specific percentiles of aPHV. Differences in trajectories for a one-year later aPHV are reported in tables. The effect of aPHV on SBP trajectories before puberty served as a negative control analysis. Any observed associations of aPHV with SBP before puberty cannot be caused by aPHV and are likely explained by confounding, particularly by adiposity, and/or possibly shared genetic architecture between SBP and puberty timing.

As is standard for inclusion of confounders in multilevel models, confounders were included as interactions with the intercept and each linear spline period. To account for the confounding effect of pre-pubertal adiposity gain, individual-specific residuals derived from multilevel models of BMI from one up to nine years of age were included as interactions with the intercept and each linear spline period. Details on multilevel models of BMI are provided in the Supplementary Material eMethods 3 and have been published previously (33). Analyses were performed with and without adjustment for confounders.

#### Additional and sensitivity analyses

We performed a number of additional and sensitivity analyses to examine generalisability, the potential for selection bias and the robustness of our findings to the number and timing of SBP measures and the pubertal age modelling strategy. Further details on these analyses are provided in Supplementary Material eMethods 4.

## Results

The characteristics of participants included in analyses, by sex, are shown in Table 1. Similar socio-demographic characteristics were observed for females and males. Mean aPHV was 11.7 years (standard deviation (SD) =0.8) for females and 13.6 years (SD=0.9) for males. Mothers of participants included in the analysis were more likely to be married, have higher household social class, higher education, higher partner education, lower prevalence of smoking during pregnancy, lower parity and higher maternal age compared with mothers of participants excluded due to missing exposure, outcome or confounder data (eTable 4). However, aPHV and SBP were similar between included and excluded participants (eTable 4).

**Table 1.** Characteristics of ALSPAC participants included in the analysis, by sex

|  | **Females**<br>**N=2,139** | **Males**<br>**N=1,923** |
| --- | --- | --- |
|  | **n (%)** | **n (%)** |
| **Maternal marital status** |  |  |
| Never married | 261(12.2) | 182(9.5) |
| Widowed\Divorced\ Separated | 86(4.0) | 85(4.4) |
| 1<sup>st</sup> Marriage | 1666(77.9) | 1520(79.0) |
| Marriage 2 or 3 | 126(5.9) | 137(7.1) |
| **Household social class †** |  |  |
| Professional | 373(17.4) | 381(19.8) |
| Managerial & Technical | 992(46.4) | 919(47.7) |
| Non-Manual | 510(23.8) | 431(22.4) |
| Manual | 191(8.9) | 134(7.0) |
| Part Skilled & Unskilled | 73(3.4) | 58(3.0) |
| **Maternal education** |  |  |
| Less than O level | 350(16.4) | 295(15.3) |
| O level | 764(35.7) | 673(35.0) |
| A level | 608(28.4) | 579(30.1) |
| Degree or above | 417(19.5) | 376(19.6) |
| **Mother's Partner's highest educational qualification** |  |  |
| Less than O level | 543(25.4) | 413(21.5) |
| O level | 454(21.2) | 426(22.2) |
| A level | 628(29.3) | 557(28.9) |
| Degree or Above | 514(24.0) | 527(27.4) |
| **Maternal smoking during pregnancy** |  |  |
| Yes | 362(16.9) | 315(16.4) |
| No | 1777(83.1) | 1608(83.6) |
| **Parity** |  |  |
| 0 | 1039(48.6) | 941(48.9) |
| 1 | 784(36.7) | 672(34.9) |
| 2+ | 316(14.8) | 310(16.1) |
| **Breastfeeding** |  |  |
| Exclusive | 829(38.7) | 685(35.6) |
| Non-exclusive | 982(45.9) | 990(51.5) |
| Never | 328(15.3) | 248(12.9) |
|  | **Mean (SD)** | **Mean (SD)** |
| **Gestational age (weeks)** | 39.6(1.5) | 39(1.8) |
| **Birth weight (g)** | 3399.8(475.4) | 3489(554.8) |
| **Maternal pre-pregnancy BMI (kg/m<sup>2</sup>)** | 22.7(3.4) | 23(3.4) |
| **Maternal age at delivery (years)** | 29.4(4.3) | 29.8(4.4) |
| **Age at puberty** | 11.7(0.8) | 13.6(0.9) |

† Household social class was measured as the highest of the mother's or her partner's occupational social class using data on job title and details of occupation collected about the mother and her partner from the mother's questionnaire at 32 weeks gestation. Social class was derived using the standard occupational classification (SOC) codes developed by the United Kingdom Office of Population Census and Surveys and classified as I professional, II managerial and technical, III non-manual, IV manual, and V part skilled occupations and unskilled occupations.

### Change in SBP before and after puberty

Mean adjusted trajectories of SBP before and after puberty in females and males at the median aPHV are presented in Figure 1. In adjusted models, females and males had similar SBP at age 3 years (Figure 1, Table 2). At puberty (median age 13.6 years in males and 11.7 years in females), males had a 10.19mmHg (95% Confidence Interval (CI): 6.80, 13.57) higher SBP compared with females (Supplementary eTable 5). By 24 years, this difference increased to 11.43mmHg (95% CI: 7.22, 15.63). Higher SBP at puberty in males appeared to be attributable to steep increases in SBP in males in the three years before puberty (Table 2). Mean adjusted SBP trajectories for females and males of similar aPHV (age 12.8 years in females [90th percentile] and 12.4 years in males [10th percentile]) are shown in Supplementary eFigure 2; at puberty males had a 5.75mmHg (95% CI: 2.30, 9.20) higher SBP compared with females; this difference increased to 10.83mmHg (95% CI: 6.41, 15.25) higher SBP in males compared with females at 24 years of age (Supplementary eTable 5).

**Figure 1.**
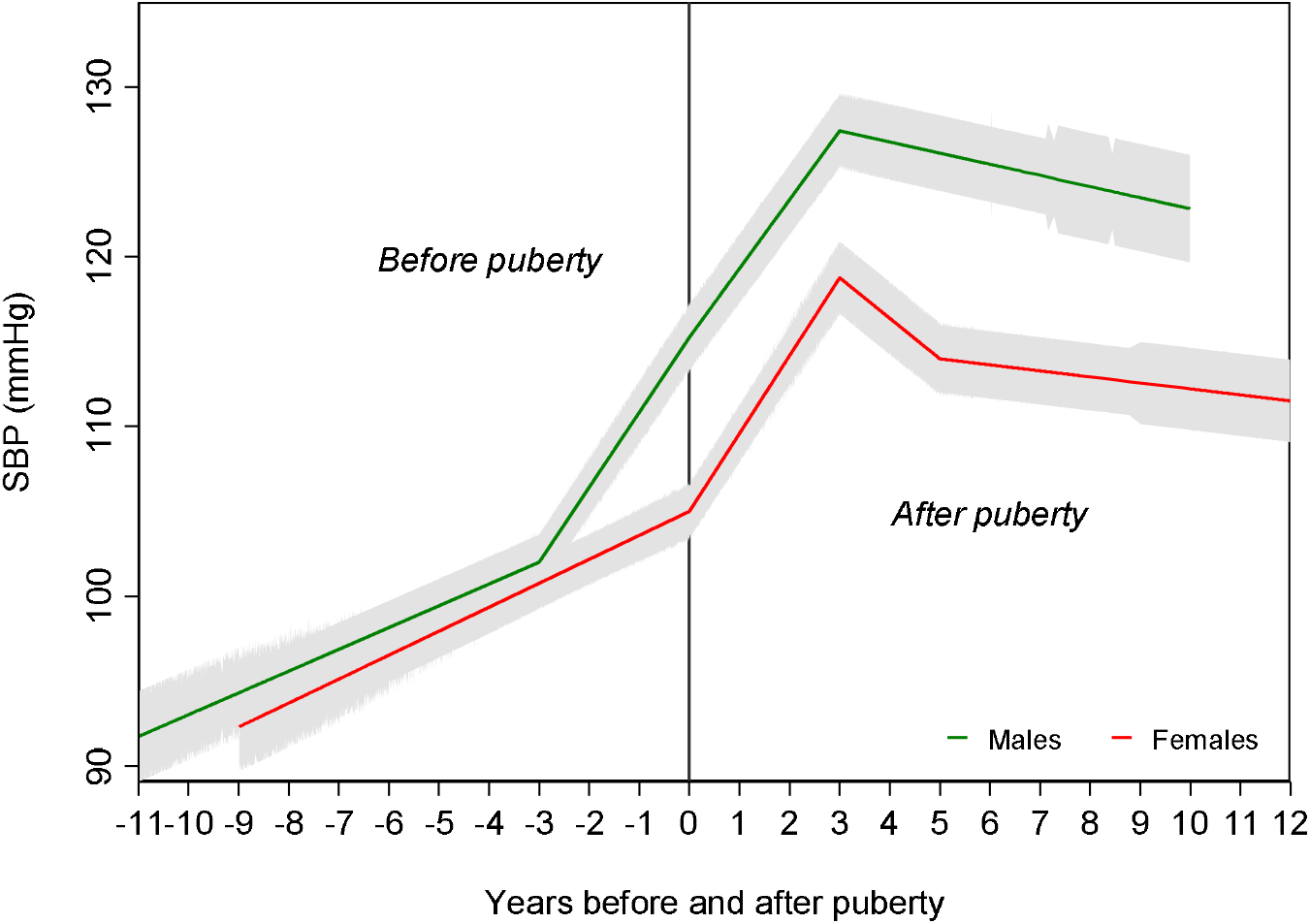
Mean adjusted trajectories of SBP in females and males before and after puberty from multilevel models based on pubertal age Age at peak height velocity is normally distributed and median is equal to mean. Models are adjusted for birth weight, gestational age, maternal education, parity, maternal smoking during pregnancy, maternal age, maternal pre-pregnancy BMI, household social class, marital status, partner education, breastfeeding, BMI residuals of offspring.

**Figure 2.**
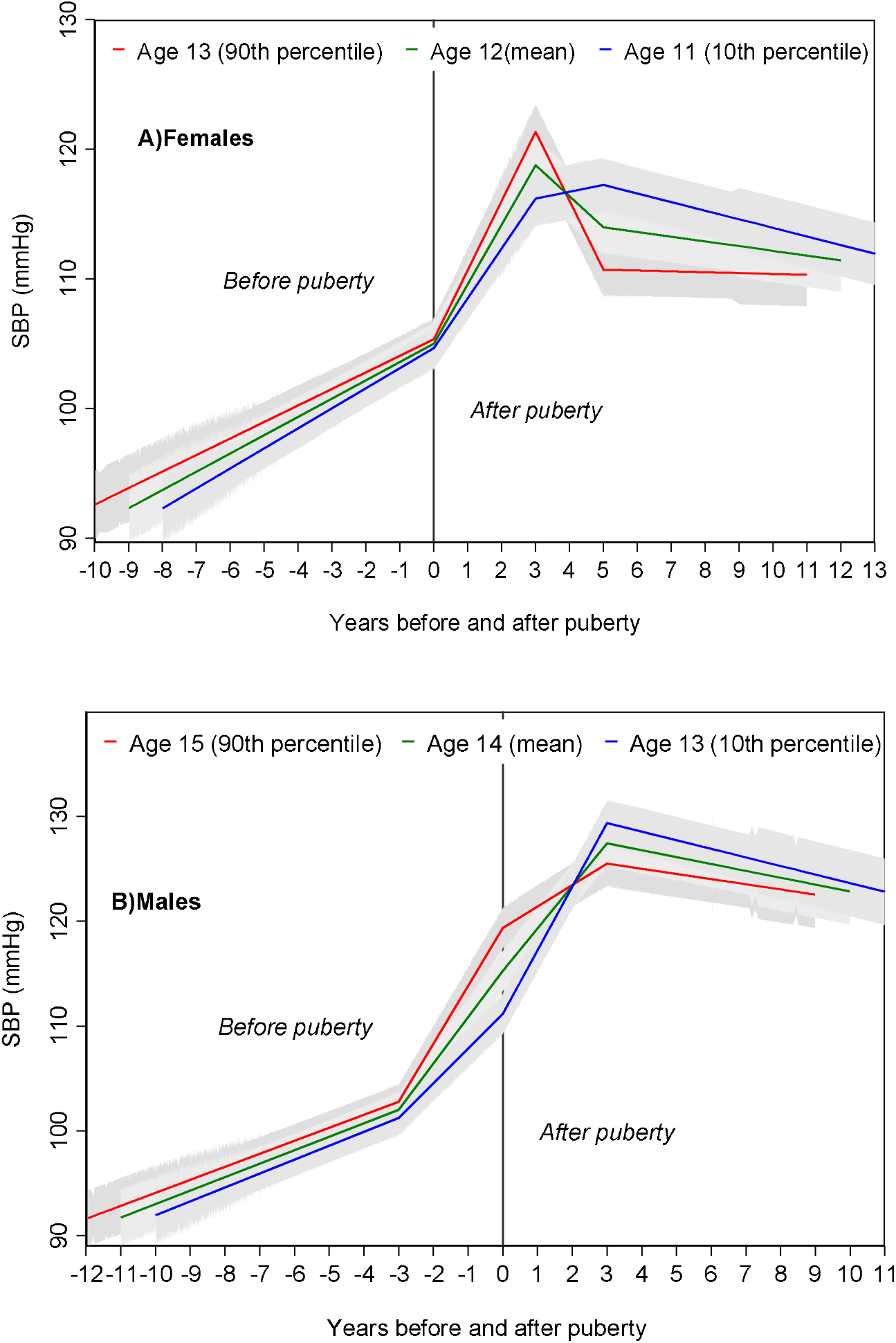
Mean adjusted trajectories of SBP in females and males for the 10^th^, median and 90^th^ sex-specific percentiles of age at peak height velocity from multilevel models based on pubertal age. Ages presented are rounded for ease of interpretation. Exact ages are 12.8y, 11.7y and 10.7y for females and 14.7y, 13.6y and 12.4y for males. Age at peak height velocity is normally distributed and median is equal to mean. Models are adjusted for birth weight, gestational age, maternal education, parity, maternal smoking during pregnancy, maternal age, maternal pre-pregnancy BMI, household social class, marital status, partner education, breastfeeding, BMI residuals of offspring.

**Table 2.** Unadjusted and adjusted mean trajectory and mean difference in trajectory of SBP per year later age at peak height velocity, from pubertal age multilevel models

|  | Unadjusted |  | Adjusted |  |
| --- | --- | --- | --- | --- |
|  | Mean trajectory (95% CI) of SBP<sup>a</sup> | Mean difference (95% CI) in SBP per year later aPHV | Mean trajectory (95% CI) of SBP<sup>a</sup> | Mean difference (95% CI) in SBP per year later aPHV |
| **Females** |  |  |  |  |
| SBP at 3 years of age (mmHg)<sup>b</sup> | 91.06 (90.47,91.66) | 1.36 (0.62,2.09) | 92.40 (89.12,95.68) | -0.13 (-1.04,0.77) |
| Change in SBP before puberty (mmHg/y) | 1.62 (1.54,1.70) | -0.09 (-0.19,-0.004) | 1.40 (0.95,1.85) | -0.12 (-0.22,-0.02) |
| SBP at puberty (mmHg) | 105.64 (105.29,105.98) | -0.69 (-1.10,-0.27) | 105.00 (103.03,106.97) | 0.31 (-0.13,0.75) |
| Change up to 3 years after puberty (mmHg/y) | 5.00 (4.80,5.21) | 0.75 (0.52,0.99) | 4.59 (3.38,5.80) | 0.68 (0.42,0.94) |
| Change 3-5 years after puberty (mmHg/y) | -2.85 (-3.17,-2.52) | -2.72 (-3.09,-2.36) | -2.36 (-4.28,-0.44) | -2.65 (-3.06,-2.24) |
| Change 5 years after puberty to end of follow-up (mmHg/y) | -0.44 (-0.51,-0.37) | 0.28 (0.21,0.37) | -0.36 (-0.79,0.07) | 0.26 (0.16,0.36) |
| SBP at age 24 years (mmHg)<sup>b</sup> | 111.87 (111.42,112.32) | -1.73 (-2.22,-1.24) | 111.54 (108.90,114.19) | -1.05 (-1.73,-0.36) |
| **Males** |  |  |  |  |
| SBP at 3 years of age (mmHg)<sup>b</sup> | 91.51 (90.87,92.15) | 0.65 (-0.05,1.35) | 91.85 (88.27,95.42) | 0.20 (-0.69,1.09) |
| Change in SBP up to 3 years before puberty (mmHg/y) | 1.32 (1.22,1.41) | -0.08 (-0.17,0.02) | 1.28 (0.74,1.82) | -0.04 (-0.14,0.06) |
| Change from 3 years before to puberty (mmHg/y) | 4.30 (4.13,4.48) | 1.07 (0.88,1.25) | 4.36 (3.32,5.41) | 1.01 (0.82,1.20) |
| SBP at puberty (mmHg) | 114.96 (114.49,115.44) | 3.16 (2.67,3.66) | 115.18 (112.43,117.93) | 3.72 (3.21,4.23) |
| Change up to 3 years after puberty (mmHg/y) | 4.02 (3.81,4.23) | -1.84 (-2.07,-1.62) | 4.05 (2.85,5.25) | -1.83 (-2.07,-1.59) |
| Change 3 years after puberty to end of follow-up (mmHg/y) | -0.57 (-0.66,-0.47) | 0.17 (0.06,0.27) | -0.62 (-1.14,-0.11) | 0.15 (0.04,0.26) |
| SBP at age 24 years (mmHg)<sup>b</sup> | 123.08 (122.50,123.65) | -0.80 (-1.36,-0.24) | 122.97 (119.70,126.24) | -0.25 (-1.03,0.54) |

<sup>a</sup> Mean trajectory is centred on the sex-specific mean of age at peak height velocity for each sex (age ~11.7 for females and age ~13.6 for males).
<sup>b</sup> Estimated using the intercept (SBP at puberty) and rates of change per year spent in each spline period before or after puberty.
Adjusted for birth weight, gestational age, maternal education, parity, maternal smoking during pregnancy, maternal age, maternal pre-pregnancy BMI, household social class, marital status, partner education, breastfeeding, BMI residuals of offspring.

### Puberty timing and SBP

#### Females

Mean adjusted female trajectories of SBP for the 10th (age 11), 50th (age 12) and 90th (age 13) percentiles of aPHV are presented in Figure 2. In adjusted models, there was no evidence of an association between a one-year later aPHV and SBP at 3 years of age (difference; −0.13mmHg; 95%CI: −1.04, 0.77) or SBP at puberty (difference; 0.31mmHg; 95% CI: −0.13, 0.75) (Table 2). A one-year later aPHV was associated with faster increases in SBP in the three years’ post-puberty and faster decreases in SBP from three to five years after puberty. From five years after puberty to the end of follow-up, a one-year later aPHV was associated with a 0.26mmHg (95% CI: 0.16, 0.36) per year slower decrease in SBP. By age 24 years, a one-year later aPHV was associated with a 1.05mmHg lower SBP (95% CI: −1.73, −0.36).

#### Males

Mean adjusted male trajectories of SBP for the 10th (age 13), 50th (age 14) and 90th (age 15) percentiles of aPHV are presented in Figure 2. Similar to females, in adjusted models there was no evidence of an association between aPHV and SBP at 3 years of age (difference; 0.20mmHg; 95% CI: - 0.69, 1.09) or in rates of change in SBP from 3 years of age to three years’ pre-puberty (difference; - 0.04mmHg per year; 95% CI: -.014, 0.06). In the three years before puberty, a one-year later aPHV was associated with a 1.01mmHg (95% CI: 0.82, 1.20) faster increase in SBP per year. At puberty, a one-year later aPHV was associated with 3.72mmHg (95% CI: 3.21, 4.23) higher SBP. In the three years after puberty, a one-year later aPHV was associated with 1.83mmHg (95% CI: −2.07, −1.59) slower increases per year and 0.15mmHg (95% CI: 0.04, 0.26) slower decreases per year in the period from three years’ post puberty to the end of follow-up. By 24 years of age, there was no evidence of a difference in SPB per year later aPHV (difference; −0.20mmHg; 95% CI: −0.69, 1.09).

#### Additional and sensitivity analyses

When analyses were conducted in the full sample of participants rather than those with complete confounder data, results were comparable (Supplementary eTable 6) as were results from the inverse probability weighted analyses (Supplementary eTable 7). Adjusting for DXA fat mass at age 9 years also resulted in similar results (Supplementary eTable 8). Results were not appreciably different when analyses were restricted to participants with at least one measure of SBP before and one measure after aPHV, or to participants with at least five measurements of SBP (Supplementary eTable 9 & 10). Results were also similar in chronological age-based models (Supplementary eTable 11).

## Discussion

In this prospective cohort study, the largest to date with an objective height-based measure of puberty timing and repeat assessments of SBP from 3 to 24 years of age, we aimed to better understand the role of puberty and its timing in sex-specific trajectories of SBP across the early life course. Our findings suggest that a large proportion of the sex difference in SBP in early adulthood is accrued before puberty with the remainder arising in the five-year period post-puberty. These findings suggest that prevention of sex differences in SBP in adulthood may benefit from a life course approach starting before puberty. Our results on puberty timing and SBP trajectories before and after puberty demonstrated no strong evidence of associations suggesting that puberty timing itself is unlikely to impact SBP in adulthood.

### Comparison with other studies

Previous life course analyses of SBP trajectories document a maximum sex difference at age 26 years with higher SBP in males compared with females (6). Our findings suggest that a large proportion of this sex difference is established before puberty with the remainder accruing in the five-year period post-puberty, regardless of whether we compare females and males of the sex-specific median age at puberty or the same ages at puberty. These results are broadly consistent with other prospective studies (14,36). A US study (n=182) examining rates of SBP change before and after puberty, defined using peak growth velocity, showed that SBP was higher in males compared with females at any given age from 5 to 25 years (36). Similar to our findings, males had nearly 8 mmHg higher SBP at puberty compared with females and rates of change in SBP were more pronounced in males with larger increases observed around the pubertal growth period. This is also consistent with previous studies documenting increasing SBP in males during adolescence compared with females (11,13).

Our findings suggested small and relatively transient associations of aPHV with SBP trajectories post puberty. By age 24 years, and after adjustment for early childhood BMI, aPHV was associated with only small differences in SBP in females and no differences in males. A Mendelian randomisation study also conducted in ALSPAC (n=3,611) found no strong evidence of associations between puberty timing (measured using reported age at menarche or voice breaking) and SBP at 18 years of age in either females or males, after adjusting for BMI measured at age 8 years (20). Results were similar to our findings with overlapping confidence intervals between the estimates in both studies. Our findings build on this evidence using an objective measure of puberty timing to reduce measurement error and improve consistency of measurement between females and males. Furthermore, using measures of height to estimate puberty timing increased both the sample size and minimised the potential for selection bias in our study compared to relying on self-report puberty questionnaires with only modest response rates. Our findings are consistent with a number of other previous studies which also demonstrated slightly lower SBP in females with later puberty timing (16,37,38). For instance, a longitudinal analysis of 391 females between the ages of 8 and 21 years in Finland showed a 1.24mmHg lower SBP per year later age at menarche (37). Our findings are also comparable with a recent sibling analysis in the Scottish Family Health Study (n=7,770) that found that later menarche was associated with a lower SBP in adulthood of a similar magnitude (38). In addition, a recent systematic review and meta-analysis of eight studies found lower SBP among women with later menarche, though confidence intervals spanned the null value (17). However, the association did strengthen when limited to high quality studies suggesting that methodological issues including heterogeneity in the definition of early menarche and small sample sizes influenced the observed association. In contrast to our findings, two studies from a British birth cohort showed some evidence of lower SBP in males late to puberty but no association in females at ages 53 and 60-54 (18,19). Measurement error may have influenced the results observed in females with puberty timing measured using mothers’ reports of age at menarche or self-report age at menarche collected when women were 48 years old while, in males, physical examinations at 15 years of age were used to categorise participants into groups of maturity stages.

### Strengths and limitations

The main strengths of our study include its prospective design, relatively large sample size, availability of repeated SBP measures from childhood to early adulthood and use of an objective measure of puberty timing (aPHV) in both sexes. Childhood adiposity is an important confounder of the association between puberty timing and SBP (20,39). To account for this, we used individual-level residual estimates from multilevel models of repeated measures of BMI from one up to nine years of age for adjustment, reducing likelihood of residual confounding by early childhood weight gain in our analysis. There are also a number of limitations. Participants excluded from the analysis due to missing data or attrition from the cohort were more socially disadvantaged than those included in our analysis leading to potential selection bias and generalisability issues. However, we aimed to minimise potential selection bias by including all participants with at least one measurement of height from 5 to <10 years, 10 to <15 years and 15 to 20 years to estimate aPHV and at least one measure of SBP from age 3 to 24 years for estimation of SBP trajectories. In addition, though some socio-demographic characteristics differed between included and excluded participants, aPHV and SBP were similarly distributed, thus minimising the impact of selection bias driven by missing exposure and outcome data in our analysis. Results from weighted sensitivity analyses and analyses with and without selection on complete confounder data were highly similar to the main findings, further indicating a low likelihood of selection bias driven by missing confounder data. Finally, the majority of our cohort were of white European ethnicity. Therefore, our findings may not be generalizable to non-white ethnicities.

## Conclusion

A large proportion of the higher SBP observed in males compared to females in early adulthood is accrued before puberty. Prevention of high SBP in adult males may therefore benefit from a life course approach starting from before puberty. Interventions targeting puberty timing are unlikely to greatly influence SBP in females and males in early adulthood.

## Data Availability

Individual-level ALSPAC data are available following an application. This process of managed access is detailed at www.bristol.ac.uk/alspac/researchers/access. Cohort details and data descriptions for ALSPAC are publicly available at the same web address.

## Acknowledgements

We are extremely grateful to all the families who took part in this study, the midwives for their help in recruiting them, and the whole ALSPAC team, which includes interviewers, computer and laboratory technicians, clerical workers, research scientists, volunteers, managers, receptionists, and nurses.

