## Supplementary Material for "Puberty timing and sex-specific trajectories of systolic blood pressure: a prospective cohort study"

**Contents**

**eMethods 1** Details of deriving age at peak height velocity

**eTable 1** Details of height measures available for deriving aPHV

**eFigure 1** Mean growth curve (black line) and velocity (blue dashed line) plots estimated by SITAR for females and males.

**eMethods 2** Details on measurement of blood pressure

**eMethods 3** Details of model selection

**eTable 2** Model details for SBP trajectories modelled using pubertal age, by sex and sex-specific quartiles of pubertal age

**eTable 3** Results from likelihood ratio test examining linearity of association between age at peak height velocity and SBP at each age by sex

**eMethods 4** Additional and sensitivity analyses

**eTable 4** Characteristics at birth of the mothers of children included in models compared with those excluded due to missing exposure, outcome or co-variate data

**eFigure 2** Mean adjusted trajectories of SBP in females and males before and after puberty from multilevel models based on pubertal age of 13

**eTable 5** Adjusted mean SBP in females and males and mean difference in SBP at age 3 years, puberty and age 24 years from pubertal age multilevel models

**eTable 6** Unadjusted mean trajectory and mean difference in trajectory of SBP per year later age at peak height velocity, from pubertal age multilevel models including all participants with data on aPHV and at least one measure of SBP from 3 to 24 years

**eTable 7** Adjusted mean trajectory and mean difference in trajectory of SBP per year later age at peak height velocity, from weighted pubertal age multilevel models

**eTable 8** Adjusted mean trajectory and mean difference in trajectory of SBP per year later age at peak height velocity, from pubertal age multilevel models – including adjustment for fat mass at age 9 years

**eTable 9** Adjusted mean trajectory and mean difference in trajectory of SBP per year later age at peak height velocity, from pubertal age multilevel models restricted to participants with at least one measure before and one measure after puberty

**eTable 10** Adjusted mean trajectory and mean difference in trajectory of SBP per year later age at peak height velocity, from pubertal age multilevel models restricted to participants with more than five measures of SBP

**eTable 11** Adjusted mean trajectory and mean difference in trajectory of SBP per year later age at peak height velocity, from chronological age multilevel models

**eMethods 1 Details of deriving age at peak height velocity**

*Height measures included in the analysis*

Height data from questionnaires and health records were excluded. Only data measured at clinic assessments carried out after age 5 years were included in the analysis. A Child in Focus (CIF) clinic measured height using a Leicester Height Measure on a 10% sub-sample of participants measured at age five. From 7 years onwards, standing height was measured to the last complete millimetre using the Harpenden Stadiometer at clinics carried out at ages 7, 9, 10, 11, 13, 14, 15 and 18 years. Data were further restricted to only include individuals with at least one measurement of height from 5 to <10 years, 10 to < 15 years and 15 to 20 years. The final dataset for analysis included 20,849 height measurements for 2,688 boys and 24,216 measurements for 3,019 girls. The number of height measures for females and males is shown in eTable 1. Further details of how age at peak height velocity (aPHV) was derived are described elsewhere (1).

*Analysis deriving aPHV*

Available height measures were analysed for females and males separately using Superimposition by Translation and Rotation (SITAR) growth curve analysis with five degrees of freedom (1,2). This method is a validated method of deriving aPHV and is described elsewhere in detail (2,3). aPHV was defined as the age when the first derivative of the mean curve, plotted as height versus age, was maximal. After fitting the initial model, the data were checked and points with velocity exceeding four standard deviations (SDs) and standardized residuals exceeding three in absolute value were removed. The model explained 98.5% of variance in males and 98.7% in females. Mean growth curve and velocity plots for females and males are shown in eFigure 1.

**eTable 1 Details of height measures available for deriving aPHV**

| **Males** |  |  | **Females** |  |  |
| --- | --- | --- | --- | --- | --- |
| **No. of height measurements per individual** | **Number of individuals** | **%** | **No. of height measurements per individual** | **Number of individuals** | **%** |
| 1 | 8 | 0.3 | N/A |  |  |
| 2 | 24 | 0.89 | 2 | 18 | 0.6 |
| 3 | 26 | 0.97 | 3 | 31 | 1.03 |
| 4 | 70 | 2.6 | 4 | 60 | 1.99 |
| 5 | 120 | 4.5 | 5 | 100 | 3.31 |
| 6 | 219 | 8.2 | 6 | 192 | 6.36 |
| 7 | 432 | 16.1 | 7 | 355 | 11.8 |
| 8 | 638 | 23.7 | 8 | 826 | 27.4 |
| 9 | 1117 | 41.6 | 9 | 1268 | 42 |
| 10 | 34 | 1.26 | 10 | 169 | 5.6 |
| Total | 2688 | 100% | Total | 3019 | 100% |

**eFigure 1 Mean growth curve (black line) and velocity (blue dashed-line) plots estimated by SITAR for females and males.**

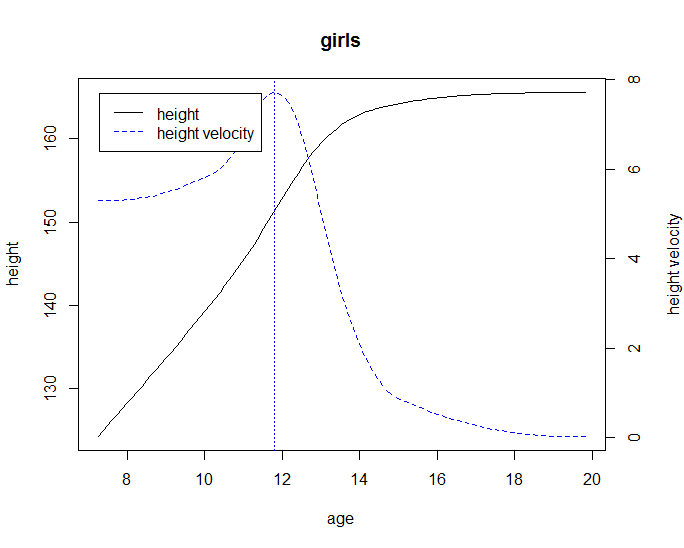

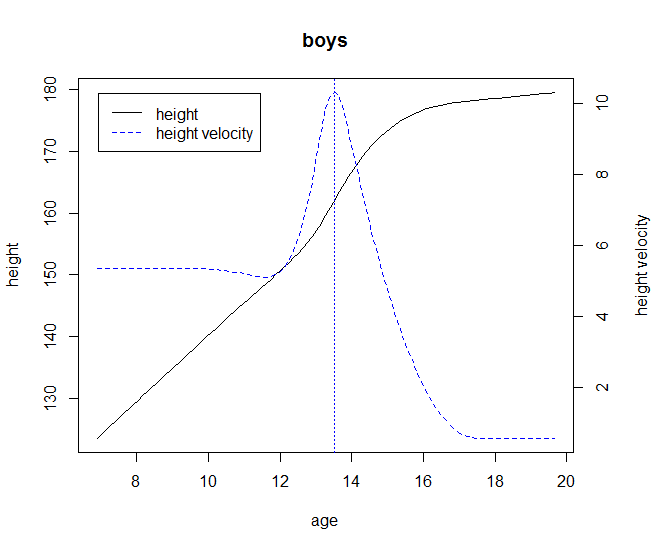

Vertical dotted line represents age at peak height velocity.

**eMethods 2 Details on measurement of blood pressure**

A Dinamap 9300 Vital Signs Monitor A was used to measure blood pressure at the 3, 4 and 5 year clinics. A Dinamap 9301 Vital Signs Monitor (Morton Medical, London) was used at the 7, 9, and 11-year clinics; an Omron MI-5 was used at the 10-year clinic; and an Omron IntelliSense M6 (Omron Healthcare, Kyoto, Japan) was used at the 15- and 18-year clinics. An Omron M6 upper arm blood pressure monitor was used at the 24-year clinic.

**eMethods 3 Details of model selection**

Systolic Blood Pressure (SBP) was measured on ten occasions between 3 and 24 years. Values of SBP four SDs greater than or less than the mean were excluded from the analysis. We included all participants with at least one measure of SBP in each multilevel model, under a missing at random (MAR), to minimise selection bias. The observations of participants who reported being pregnant at the 18-year and 24-year clinics were excluded from the multilevel models at that time point only. We modelled sex-specific change over time in SBP according to pubertal age to better understand the association of age at peak height velocity with change in SBP during childhood and adolescence. In both models, linear splines were used to examine change in SBP (4). aPHV was normally distributed in both sexes. Linearity of associations of aPHV and change in SBP was examined by comparing the model fit of regressions of SBP on aPHV, with aPHV treated as a continuous exposure and as a categorical exposure (fourths of aPHV). Linearity was formally tested using a likelihood ratio test (see eTable 3). Prior to analysis, aPHV was centred on the sex-specific mean of aPHV for females and males.

*Models based on pubertal age*

Models examining change in SBP according to pubertal age were modelled de novo for this paper. We examined observed data at each age by sex to examine whether the shape of change over time was similar or different between quartiles of pubertal age. Shape of change over time across quartiles of pubertal age differed for females and males. Therefore, based on the observed data, we examined the fit of two models with four periods of change (one pre- and three post-puberty) and another model with five periods of change (two pre- and three post-puberty) separately in females and males. The model with the best fit in both females and males across each quartile of pubertal age was a four-spline model allowing for four periods of change; one pre-pubertal period (from three years before puberty to puberty) and three post-pubertal periods (from puberty to three years after puberty, three to five years after puberty and from five years after puberty to the end of follow-up) in females and two pre-pubertal periods (from three years to three years before puberty, from three years before puberty to puberty) and two post-pubertal periods (from puberty to three years after puberty, and from three years after puberty to the end of follow-up) in males.

The models for took the form of SBP_ij_ = β_0_ + u_0j_ + (β_1_+ u_1j_ )s_ij1_ + (β_2_+ u_2j_ )s_ij2_ + (β_3_ + u_3j )_s_ij3_ + (β_4_ + u_4j )_s_ij4_ + e_ij_ where for person j at measurement occasion i; β_0_ represents the fixed effect coefficient for the average intercept, β_1_ represents fixed effect coefficients for the first average linear slope, β_2_ represents fixed effect coefficients for the second average linear slope, β_3_ represents the fixed effect coefficients for the third average linear slope, β_4_ represents the fixed effect coefficients for the final average linear slope, u_0j_ to u_3j_ indicate person-specific random effects for the intercept and slopes respectively, and e_ij_ represents the occasion-specific residuals or measurement error which was allowed to vary with age. The covariance between the final two spline periods was set to zero to improve model convergence.

*Measurement of confounders adjusted included in main analyses*

Birthweight was extracted from medical records. Gestational age at birth was estimated from clinical records. A questionnaire at 32 weeks gestation asked mothers to report their educational attainment, which was categorized as below O-Level (Ordinary Level; exams taken in different subjects usually at age 15-16 at the completion of legally required school attendance, equivalent to today’s UK General Certificate of Secondary Education), O-Level only, A-Level (Advanced-Level; exams taken in different subjects usually at age 18), or university degree or above. Parity was defined as the number of previous pregnancies that had resulted in a live- or still-born infant collected at 18 weeks gestation. Smoking in the first trimester of pregnancy was self-reported by mothers at 18 weeks gestation; responses to smoking any tobacco (cigarettes, cigars, pipes, or other) were grouped as follows: no smoking, <10 per day, 10-19 per day or greater than 19 per day. Maternal age was reported in the mother’s antenatal questionnaires. Maternal height and weight were self-reported from the questionnaire administered at 12 weeks gestation; these were used to calculate maternal BMI. Household social class was measured as the highest of the mother’s or her partner’s occupational social class using data on job title and details of occupation collected about the mother and her partner from the mother’s questionnaire at 32 weeks gestation. Social class was derived using the standard occupational classification (SOC) codes developed by the United Kingdom Office of Population Census and Surveys and classified as I professional, II managerial and technical, IIINM non-manual, IIIM manual, and IV&V part skilled occupations and unskilled occupations. Marital status was obtained from antenatal questionnaires and classified as never married, widowed, divorced, separated, first marriage, marriage two or three. A questionnaire at 32 weeks gestation asked partners to report their educational attainment, which was categorized as below O-Level (Ordinary Level; exams taken in different subjects usually at age 15-16y at the completion of legally required school attendance, equivalent to today’s UK General Certificate of Secondary Education), O-Level only, A-Level (Advanced-Level; exams taken in different subjects usually at age 18), or university degree or above. Breastfeeding information used here was collected via questionnaires administered at 4 weeks, 6 months and 15 months.

*Body Mass Index*

From 1 to 5 years, measures were available from routine child health clinics for most children and extracted from health visitor records, which form part of standard child care in the UK. Data were also available from research clinic measurements on a random 10% subsample of the cohort. All cohort members were invited to research clinics from age 7 onwards. Across all ages parent-reported measures were available.

At the clinics, crown-heel length for children aged four to 25 months was measured using a Harpenden Neonatometer and from 25 months onwards standing height was measured using a Leicester Height Measure; weight was measured using Fereday 100kg combined scale (four-month clinic), Soenhle scale or Seca scale model 724 (eight-month clinic), Seca 724 or Seca 835 (12-month clinic), Seca 835 (18 months onwards). From age 7 years, all children were invited to annual clinics, at which standing height was measured to the last complete mm using the Harpenden Stadiometer and weight was measured to the nearest 0.1kg using the Tanita Body Fat Analyser (Model TBF 305).

BMI has been modelled previously using fractional polynomials and is described elsewhere. Briefly, BMI was log transformed due to skewness of the data and fractional polynomials were used where age was raised to various combinations of powers (each of the following single powers, plus each combination of two powers: 0.5, 1, 2, 3, -0.5, -1, -2, natural log), from which we selected the best fitting curve (the one with the lowest likelihood value). The resulting curve contained three age terms including log age, log age* age and log age *age^2. To account for the likely reduced accuracy of parent-reported measurements, a binary indicator of measurement source (research clinic or health records versus parent-report) was included as a fixed effect. The variance of measurement occasion-level residuals (the differences between observed and predicted measurements) was allowed to vary with age for log BMI. The model took the form of: log BMIij = (β0+u0j+e0ij) + (β1+u1j)(ln(age)ij) + (β2+u2j)(age*ln(age)ij) + (β3+u3j)(age2*ln(age)ij) + (β8+e1ij)(measurement_sourceij) + eij(age_monthsij) where for person j at measurement occasion i; β’s represent fixed effect coefficients, u0j to u3j indicate person-specific random effects for the intercept and linear, quadratic and cubic age terms respectively, and e1 represents the occasion-specific residuals or measurement error which was allowed to vary with age and according to measurement source.

Individual-specific residuals were derived from fitting these multilevel models of weight and height from one up to nine years of age, dropping all measurements before 12 months and after 108 months, fitted using the statistical software package MLWiN version 3.04.

|  | No of contributing individuals | | Assessment of model fit | | | | |
| --- | --- | --- | --- | --- | --- | --- | --- |
|  | Total number of observations | Number of individuals with 1 measure | Mean observed (SD), SBP | Mean predicted (SD), SBP | Mean difference (observed – predicted), SBP | 95% level of agreement between observed and predicted, SBP | |
| Females |  |  |  |  |  | |  |
| Overall | 15509 | 2139 |  |  |  | |  |
| 1^st^ quartile (9.0-11.2) | 3886 | 537 | 109.90 (11.76) | 109.76 (8.43) | 0.14 | | -12.96 to 13.24 |
| 2^nd^ quartile (11.2-11.7) | 3875 | 533 | 107.82 (11.58) | 107.80 (8.61) | 0.02 | | -12.19 to 12.22 |
| 3^rd^ quartile (11.7-12.3) | 3873 | 540 | 106.73 (11.54) | 106.79 (8.88) | -0.06 | | -11.64 to 11.51 |
| 4^th^ quartile (12.3-14.6) | 3875 | 529 | 105.13 (11.53) | 105.22 (9.08) | -0.10 | | -12.11 to 11.92 |
| Males |  |  |  |  |  | |  |
| Overall | 13663 | 1923 |  |  |  | |  |
| 1^st^ quartile (9.7-13.0) | 3419 | 475 | 111.83 (14.55) | 111.86 (12.04) | -0.03 | | -12.55 to 12.50 |
| 2^nd^ quartile (13.0-13.6) | 3415 | 482 | 109.76 (14.15) | 109.82 (11.84) | -0.06 | | -12.35 to 12.23 |
| 3^rd^ quartile (13.6-14.2) | 3414 | 480 | 108.70 (13.97) | 108.70 (11.80) | 0.007 | | -12.28 to 12.29 |
| 4^th^ quartile (14.2-17.4) | 3415 | 486 | 107.05 (13.47) | 106.97 (11.20) | 0.08 | | -12.45 to 12.62 |

**eTable 2 Model details for SBP trajectories modelled using pubertal age, by sex and sex-specific quartiles of pubertal age**

SD, Standard Deviation; SBP, Systolic Blood Pressure

**eTable 3 Results from likelihood ratio test examining linearity of association between age at peak height velocity and SBP at each age by sex**

|  | **Females** | **Males** |
| --- | --- | --- |
|  | **P value comparing models** | **P value comparing models** |
| Age 3 | 0.15 | 0.40 |
| Age 5 | 0.28 | 0.26 |
| Age 7 | 0.50 | 0.20 |
| Age 9 | 0.13 | 0.51 |
| Age 11 | 0.17 | 0.004 |
| Age 13 | 0.40 | 0.10 |
| Age 15 | 0.95 | 0.81 |
| Age 18 | 0.95 | 0.26 |
| Age 24 | 0.71 | 0.25 |

P value from likelihood ratio test comparing fit of models regressing thirds of pubertal age treated as a continuous exposure on SBP at each age to models regressing thirds of pubertal age treated as categorical exposure on SBP at each age. P>0.05 indicates the more parsimonious model (pubertal age treated as continuous exposure) is a better fit, suggesting linearity of associations of age at peak height velocity and SBP. Note that although the p value for males at age 11 indicated some departure from linearity, associations were still broadly linear. Thus, given the linearity of all other associations in females and males, age at peak height velocity was examined as a continuous exposure in our analyses.

**eMethods** **4 Additional and sensitivity analyses**

We examined the characteristics of participants included in our analysis compared with participants excluded from our analysis due to missing exposure, outcome or confounder data, using socio-demographic characteristics measured at or close to birth to better understand generalisability and the potential for selection bias. We performed weighted sensitivity analyses using inverse probability weighting to address potential selection bias. The participant level weights were estimated using logistic regression using all socio-demographic characteristics listed above as potential confounders with the addition of gender and were subsequently incorporated into the multi-level models as level two weights which adjust for the unequal probability of selection of the participants (36). We additionally performed unadjusted analyses on the sample of participants that had data on aPHV and at least one measure of SBP from 3 to 24 years; this analysis included an additional 2,517 participants excluded from our main analysis due to missing confounder data and provided insight into potential selection due to missing confounder data. To further account for the potential confounding effect of pre-pubertal adiposity, we performed analyses adjusted for fat mass which was directly measured from whole body dual energy X-ray absorptiometry (DXA) scans at nine years of age. We also performed sensitivity analyses restricting the sample to participants with at least one SBP measure before and one after aPHV and to those with a minimum of five SBP measures in total during follow-up. Finally, we explored the association between aPHV and chronological age-based trajectories of SBP using sex-stratified linear spline multilevel models with five periods of linear change (age 3-7 years, 7-12 years, 12-16 years, 16-18 years and 18-24 years) and compared results to findings from the pubertal age-based models used in our main analysis.

SBP was previously modelled according to chronological age using linear spline multi-level models from 7 to 18 years. The model for SBP is described elsewhere in detail (3,5). The knots for the model were placed at 12 and 16 resulting in three periods of change: from 7-12, 12-16 and 16-18. To model SBP from 3 to 24 years of age, we added two additional linear splines so that there were five periods of change: from 3-7, 7-12, 12-16, 16-20, 20-24. The models took the form of: SBP_ij_ = β_0_ + u_0j_ + (β_1_+ u_1j_ )s_ij1_ + (β_2_+ u_2j_ )s_ij2_ + (β_3_ + u_3j_ )s_ij3_ + (β_4_ + u_4j_ )s_ij4_ + (β_5_ + u_5j_ )s_ij5_ + e_ij_ where for person j at measurement occasion i; β_0_ represents the fixed effect coefficient for the average intercept, β_1_ to β_5_ represent fixed effect coefficients for the average linear slopes of each linear spline, s_ij_ represents the specific spline period, u_0j_ to u_5j_ indicate person-specific random effects for the intercept and slopes respectively, and e_ij_ represents the occasion-specific residuals or measurement error which was allowed to vary with age.

For this analysis, we examined whether this model was appropriate for modelling change over time within quartiles of pubertal age to ensure that model fit was good across the entire distribution of pubertal age. The association between aPHV and chronological age-based trajectories was then examined separately for females and males by including an interaction between centred sex-specific aPHV and the intercept and each spline period, providing an estimate of the difference in the average trajectory of SBP from age 3 years to 24 years, per year later aPHV. Confounders were included as interactions with both the intercept and linear slopes.

**eTable 4 Characteristics at birth of the mothers of children included in models compared with those excluded due to missing exposure, outcome or co-variate data**

|  | **Participants included**  **n= 4,062** | **Participants excluded**  **n=1,639-11,580 ^a^** | ***P* value for comparison** † |
| --- | --- | --- | --- |
|  | **n (%)** | **n (%)** |  |
| **Maternal marital status** |  |  |  |
| Never married | 443(10.9) | 2153(22.7) | <0.001 |
| Widowed/ Divorced/ Separated | 170(4.2) | 646(6.8) |  |
| 1^st^ Marriage | 3186(78.4) | 6069(64.0) |  |
| Marriage 2 or 3 | 263(6.5) | 618(6.5) |  |
| **Household social class** |  |  |  |
| Professional | 754(18.6) | 785(10.5) | <0.001 |
| Managerial & Technical | 1911(47.0) | 2917(38.9) |  |
| Non-Manual | 941(23.2) | 2006(26.7) |  |
| Manual | 325(8.0) | 1238(16.5) |  |
| Part Skilled & Unskilled | 131(3.2) | 561(7.5) |  |
| **Maternal education** |  |  |  |
| Less than O level | 645(15.9) | 3110(36.9) | <0.001 |
| O level | 1437(35.4) | 2886(34.3) |  |
| A level | 1187(29.2) | 1608(19.1) |  |
| Degree or above | 793(19.5) | 816(9.7) |  |
| **Partners highest educational qualification** |  |  |  |
| Less than O level | 956(23.5) | 3193(40.2) | <0.001 |
| O level | 880(21.7) | 1675(21.1) |  |
| A level | 1185(29.2) | 1933(24.4) |  |
| Degree or Above | 1041(25.6) | 1137(14.3) |  |
| **Maternal smoking during pregnancy** |  |  |  |
| Yes | 677(16.7) | 2289(24.8) | <0.001 |
| No | 3385(83.3) | 6937(75.2) |  |
| **Parity** |  |  |  |
| 0 | 1980(48.7) | 3889(34.3) | <0.001 |
| 1 | 1456(35.8) | 3126(27.6) |  |
| 2+ | 626(15.4) | 4317(38.1) |  |
| **Sex** |  |  |  |
| Female | 1989(47.6) | 5638(52.8) | <0.001 |
| Male | 2192(52.4) | 5031(47.2) |  |
|  | ***Mean (SD)*** | ***Mean (SD)*** | ***P* value** |
| Child gestational age at birth | 39.5(1.7) | 37.9(6.3) | <0.001 |
| Birthweight (g) | 3442(516.4) | 3356(603.7) | <0.001 |
| Maternal BMI (kg/m^2^) | 22.8(3.4) | 22.8(3.6) | 0.07 |
| Maternal age (years) | 29.6(4.4) | 27.7(4.9) | <0.001 |
| Mean age at peak height velocity (years) | 12.6(1.3) | 12.6(1.3) | 0.67 |
| SBP at age 3 years  SBP at age 15 years  SBP at age 24 years | 90.4(7.8)  122.9(10.8)  116.2(11.4) | 90.0 (7.8)  122.9(11.3)  115.4(10.9) | 0.48  0.79  0.03 |

^a^ Denominators for excluded participants in this table varies due to missing data for characteristics shown.

**eFigure 2** **Mean adjusted trajectories of SBP in females and males before and after puberty from multilevel models based on pubertal age of 13**

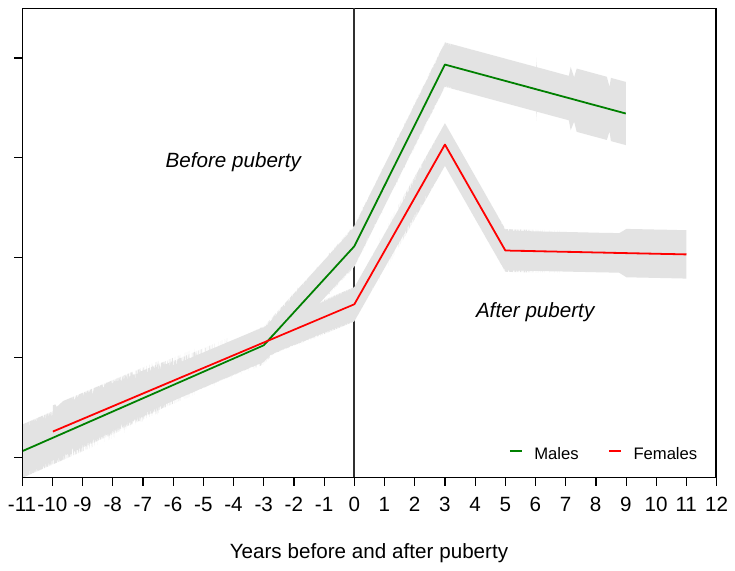

Models are adjusted for birth weight, gestational age, maternal education, parity, maternal smoking during pregnancy, maternal age, maternal pre-pregnancy BMI, household social class, marital status, partner education, breastfeeding, BMI residuals. Exact ages are 12.8 years for females and 12.4 years for males. SBP, Systolic Blood Pressure; mmHg/y, millimetres of mercury per year

**eTable 5 Adjusted mean SBP in females and males and mean difference in SBP, from pubertal age multilevel models**

|  | **Mean SBP (95% CI)^a^ in females** | **Mean SBP (95% CI)^a^ in males** | **Mean difference in SBP (95% CI)** | **P-value** |
| --- | --- | --- | --- | --- |
| **Median age at peak height velocity** |  |  |  |  |
| SBP at 3 years of age (mmHg) | 92.40 (89.12,95.68) | 91.85 (88.27,95.42) | 0.55 (-4.30,5.41) | 0.8 |
| SBP at puberty (mmHg) | 105.00 (103.02,106.97) | 115.18 (112.43,117.93) | -10.19 (-13.57,-6.80) | <0.001 |
| SBP at age 24 years (mmHg) | 111.54 (108.90,114.19) | 122.97 (119.70,126.24) | -11.43 (-15.63,-7.22) | <0.001 |
| **Age 13 years at peak height velocity** |  |  |  |  |
| SBP at 3 years of age (mmHg) | 92.69 (88.94,96.44) | 92.03 (88.82,95.24) | 0.66 (-4.28,5.59) | 0.8 |
| SBP at puberty (mmHg) | 105.33 (103.31,107.36) | 111.09 (108.29,113.88) | -5.75 (-9.20,-2.30) | <0.001 |
| SBP at age 24 years (mmHg) | 112.15 (109.76,114.54) | 122.98 (119.26,126.69) | -10.83 (-15.25,-6.41) | <0.001 |

^a^ Estimated using regression coefficients for the intercept and rates of change per year spent in each spline period.

^b^ Difference in SBP between females and males at age 3, at puberty and age 24 was estimated by calculating the mean difference between the sexes and using the pooled standard error to calculate 95% confidence intervals for the difference.

Median of age at peak height velocity is age 11.7 years for females and age 13.6 for males. Exact ages for age 13 years at peak height velocity are 12.8 years for females and 12.4 years for males.

Models are adjusted for birth weight, gestational age, maternal education, parity, maternal smoking during pregnancy, maternal age, maternal pre-pregnancy BMI, household social class, marital status, partner education, breastfeeding, BMI residuals.

SBP, Systolic Blood Pressure; CI, confidence interval; mmHg/y, millimetres of mercury per year.

**eTable 6 Unadjusted mean trajectory and mean difference in trajectory of SBP per year later age at peak height velocity, from pubertal age multilevel models including all participants with data on aPHV and at least one measure of SBP from 3 to 24 years**

|  | **Mean trajectory (95% CI) of SBP^a^** | **Mean difference (95% CI) in SBP per year later aPHV** |
| --- | --- | --- |
| **Females** |  |  |
| SBP at 3 years of age (mmHg)^b^ | 90.99 (90.46,91.51) | 1.50 (0.85,2.15) |
| Change in SBP before puberty (mmHg/y) | 1.64 (1.57,1.71) | -0.08 (-0.16,0.004) |
| SBP at puberty (mmHg) | 105.75 (105.46,106.04) | -0.63 (-0.98,-0.27) |
| Change up to 3 years after puberty (mmHg/y) | 4.91 (4.74,5.09) | 0.76 (0.56,0.97) |
| Change 3-5 years after puberty (mmHg/y) | -2.56 (-2.85,-2.27) | -2.80 (-3.13,-2.47) |
| Change 5 years after puberty to end of follow-up (mmHg/y) | -0.51 (-0.57,-0.44) | 0.32 (0.25,0.40) |
| SBP at age 24 years (mmHg) ^b^ | 111.84 (111.46,112.21) | -1.49 (-1.90,-1.08) |
| **Males** |  |  |
| SBP at 3 years of age (mmHg) ^b^ | 91.81 (91.23,92.38) | 0.65 (0.02,1.29) |
| Change in SBP up to 3 years before puberty (mmHg/y) | 1.31 (1.22,1.39) | -0.09 (-0.17,0.001) |
| Change from 3 years before to puberty (mmHg/y) | 4.23 (4.08,4.38) | 1.05 (0.89,1.20) |
| SBP at puberty (mmHg) | 114.96 (114.55,115.36) | 3.02 (2.60,3.43) |
| Change up to 3 years after puberty (mmHg/y) | 4.08 (3.90,4.26) | -1.76 (-1.95,-1.57) |
| Change 3 years after puberty to end of follow-up (mmHg/y) | -0.60 (-0.68,-0.52) | 0.14 (0.06,0.23) |
| SBP at age 24 years (mmHg) ^b^ | 122.98 (122.48,123.47) | -0.81 (-1.28,-0.33) |

^a^Mean trajectory is centred on the sex-specific median of age at peak height velocity for each sex (age ~11.7 for females and age ~13.6 for males).

^b^ Estimated using regression coefficients for the intercept and rates of change per year spent in each spline period.

SBP, Systolic Blood Pressure; CI, confidence interval; mmHg/y, millimetres of mercury per year.

**eTable 7 Adjusted mean trajectory and mean difference in trajectory of SBP per year later age at peak height velocity, from weighted pubertal age multilevel models**

|  | **Mean trajectory (95% CI) of SBP^a^** | **Mean difference (95% CI) in SBP per year later aPHV** |
| --- | --- | --- |
| **Females** |  |  |
| SBP at 3 years of age (mmHg)^b^ | 93.50 (90.14,96.85) | -0.44 (-1.44,0.57) |
| Change in SBP before puberty (mmHg/y) | 1.24 (0.79,1.69) | -0.14 (-0.26,-0.03) |
| SBP at puberty (mmHg) | 104.65 (102.67,106.64) | 0.25 (-0.22,0.72) |
| Change up to 3 years after puberty (mmHg/y) | 4.82 (3.51,6.14) | 0.76 (0.49,1.04) |
| Change 3-5 years after puberty (mmHg/y) | -2.56 (-4.61,-0.51) | -2.70 (-3.14,-2.27) |
| Change 5 years after puberty to end of follow-up (mmHg/y) | -0.38 (-0.83,0.07) | 0.24 (0.14,0.35) |
| SBP at age 24 years (mmHg) ^b^ | 111.36 (108.64,114.08) | -1.02 (-1.71,-0.32) |
| **Males** |  |  |
| SBP at 3 years of age (mmHg) ^b^ | 91.85 (88.27,95.42) | 0.20 (-0.69,1.09) |
| Change in SBP up to 3 years before puberty (mmHg/y) | 1.28 (0.74,1.82) | -0.04 (-0.14,0.06) |
| Change from 3 years before to puberty (mmHg/y) | 4.36 (3.32,5.41) | 1.01 (0.82,1.20) |
| SBP at puberty (mmHg) | 115.18 (112.43,117.93) | 3.72 (3.21,4.23) |
| Change up to 3 years after puberty (mmHg/y) | 4.05 (2.85,5.25) | -1.83 (-2.07,-1.59) |
| Change 3 years after puberty to end of follow-up (mmHg/y) | -0.62 (-1.14,-0.11) | 0.15 (0.04,0.26) |
| SBP at age 24 years (mmHg) ^b^ | 122.97 (119.70,126.24) | -0.25 (-1.03,0.54) |

^a^Mean trajectory is centred on the sex-specific median of age at peak height velocity for each sex (age ~11.7 for females and age ~13.6 for males).

^b^ Estimated using regression coefficients for the intercept and rates of change per year spent in each spline period.

Adjusted for birth weight, gestational age, maternal education, parity, maternal smoking during pregnancy, maternal age, maternal pre-pregnancy BMI, household social class, marital status, partner education, breastfeeding, BMI residuals.

SBP, Systolic Blood Pressure; CI, confidence interval; mmHg/y, millimetres of mercury per year.

**eTable 8 Adjusted mean trajectory and mean difference in trajectory of SBP per year later age at peak height velocity, from pubertal age multilevel models – including adjustment for fat mass at age 9 years**

|  | **Mean trajectory (95% CI) of SBP^a^** | **Mean difference (95% CI) in SBP per year later aPHV** |
| --- | --- | --- |
| **Females** |  |  |
| SBP at 3 years of age (mmHg)^b^ | 85.97 (82.33,89.60) | 0.16 (-0.73,1.06) |
| Change in SBP before puberty (mmHg/y) | 1.63 (1.14,2.12) | -0.11 (-0.21,-0.003) |
| SBP at puberty (mmHg) | 100.61 (98.45,102.76) | 0.40 (-0.05,0.85) |
| Change up to 3 years after puberty (mmHg/y) | 4.60 (3.31,5.89) | 0.65 (0.38,0.92) |
| Change 3-5 years after puberty (mmHg/y) | -2.86 (-4.92,-0.81) | -2.59 (-3.02,-2.17) |
| Change 5 years after puberty to end of follow-up (mmHg/y) | -0.06 (-0.53,0.40) | 0.26 (0.16,0.36) |
| SBP at age 24 years (mmHg) ^b^ | 108.22 (105.30,111.14) | -1.20 (-1.95,-0.45) |
| **Males** |  |  |
| SBP at 3 years of age (mmHg) ^b^ | 90.29 (86.39,94.19) | 0.03 (-0.90,0.95) |
| Change in SBP up to 3 years before puberty (mmHg/y) | 1.02 (0.44,1.60) | 0.001 (-0.11,0.11) |
| Change from 3 years before to puberty (mmHg/y) | 4.53 (3.43,5.64) | 1.00 (0.80,1.20) |
| SBP at puberty (mmHg) | 112.07 (109.14,115.00) | 4.01 (3.47,4.55) |
| Change up to 3 years after puberty (mmHg/y) | 4.42 (3.15,5.70) | -1.91 (-2.16,-1.65) |
| Change 3 years after puberty to end of follow-up (mmHg/y) | -0.73 (-1.29,-0.18) | 0.18 (0.06,0.29) |
| SBP at age 24 years (mmHg) ^b^ | 120.23 (116.72,123.74) | 0.10 (-0.77,0.96) |

^a^Mean trajectory is centred on the sex-specific median of age at peak height velocity for each sex (age ~11.7 for females and age ~13.6 for males).

^b^ Estimated using regression coefficients for the intercept and rates of change per year spent in each spline period.

Adjusted for birth weight, gestational age, maternal education, parity, maternal smoking during pregnancy, maternal age, maternal pre-pregnancy BMI, household social class, marital status, partner education, breastfeeding, fat mass at age nine years.

SBP, Systolic Blood Pressure; CI, confidence interval; mmHg/y, millimetres of mercury per year.

**eTable 9 Adjusted mean trajectory and mean difference in trajectory of SBP per year later age at peak height velocity, from pubertal age multilevel models restricted to participants with at least one measure before and one measure after puberty**

|  | **Mean trajectory (95% CI) of SBP^a^** | **Mean difference (95% CI) in SBP per year later aPHV** |
| --- | --- | --- |
| **Females** |  |  |
| SBP at 3 years of age (mmHg)^b^ | 92.66 (89.36,95.95) | -0.14 (-1.05,0.77) |
| Change in SBP before puberty (mmHg/y) | 1.37 (0.92,1.82) | -0.12 (-0.22,-0.02) |
| SBP at puberty (mmHg) | 104.97 (102.99,106.95) | 0.31 (-0.13,0.76) |
| Change up to 3 years after puberty (mmHg/y) | 4.65 (3.44,5.86) | 0.65 (0.39,0.91) |
| Change 3-5 years after puberty (mmHg/y) | -2.37 (-4.30,-0.45) | -2.62 (-3.03,-2.20) |
| Change 5 years after puberty to end of follow-up (mmHg/y) | -0.35 (-0.79,0.08) | 0.25 (0.16,0.35) |
| SBP at age 24 years (mmHg) ^b^ | 111.69 (109.04,114.35) | -1.10 (-1.79,-0.40) |
| **Males** |  |  |
| SBP at 3 years of age (mmHg) ^b^ | 91.69 (88.08,95.30) | 0.04 (-0.95,1.03) |
| Change in SBP up to 3 years before puberty (mmHg/y) | 1.27 (0.73,1.81) | -0.07 (-0.19,0.04) |
| Change from 3 years before to puberty (mmHg/y) | 4.43 (3.38,5.48) | 1.08 (0.88,1.29) |
| SBP at puberty (mmHg) | 115.13 (112.37,117.90) | 3.82 (3.29,4.35) |
| Change up to 3 years after puberty (mmHg/y) | 4.02 (2.82,5.22) | -1.88 (-2.12,-1.63) |
| Change 3 years after puberty to end of follow-up (mmHg/y) | -0.63 (-1.15,-0.11) | 0.15 (0.05,0.26) |
| SBP at age 24 years (mmHg) ^b^ | 122.79 (119.52,126.06) | -0.26 (-1.05,0.52) |

^a^Mean trajectory is centred on the sex-specific median of age at peak height velocity for each sex (age ~11.7 for females and age ~13.6 for males).

^b^ Estimated using regression coefficients for the intercept and rates of change per year spent in each spline period.

Adjusted for birth weight, gestational age, maternal education, parity, maternal smoking during pregnancy, maternal age, maternal pre-pregnancy BMI, household social class, marital status, partner education, breastfeeding, BMI residuals.

SBP, Systolic Blood Pressure; CI, confidence interval; mmHg/y, millimetres of mercury per year.

**eTable 10 Adjusted mean trajectory and mean difference in trajectory of SBP per year later age at peak height velocity, from pubertal age multilevel models restricted to participants with more than five measures of SBB**

|  | **Mean trajectory (95% CI) of SBP^a^** | **Mean difference (95% CI) in SBP per year later aPHV** |
| --- | --- | --- |
| **Females** |  |  |
| SBP at 3 years of age (mmHg)^b^ | 92.48 (89.09,95.87) | -0.04 (-0.97,0.89) |
| Change in SBP before puberty (mmHg/y) | 1.36 (0.90,1.83) | -0.12 (-0.22,-0.02) |
| SBP at puberty (mmHg) | 104.74 (102.66,106.83) | 0.20 (-0.26,0.66) |
| Change up to 3 years after puberty (mmHg/y) | 4.21 (2.96,5.47) | 0.67 (0.40,0.94) |
| Change 3-5 years after puberty (mmHg/y) | -1.60 (-3.61,0.40) | -2.68 (-3.11,-2.25) |
| Change 5 years after puberty to end of follow-up (mmHg/y) | -0.43 (-0.88,0.02) | 0.28 (0.18,0.38) |
| SBP at age 24 years (mmHg) ^b^ | 111.16 (108.37,113.95) | -1.06 (-1.78,-0.34) |
| **Males** |  |  |
| SBP at 3 years of age (mmHg) ^b^ | 91.64 (87.95,95.33) | 0.26 (-0.65,1.18) |
| Change in SBP up to 3 years before puberty (mmHg/y) | 1.26 (0.71,1.82) | -0.02 (-0.13,0.08) |
| Change from 3 years before to puberty (mmHg/y) | 4.38 (3.30,5.46) | 1.02 (0.82,1.22) |
| SBP at puberty (mmHg) | 114.89 (112.02,117.76) | 3.83 (3.30,4.37) |
| Change up to 3 years after puberty (mmHg/y) | 3.79 (2.55,5.03) | -1.80 (-2.05,-1.55) |
| Change 3 years after puberty to end of follow-up (mmHg/y) | -0.60 (-1.13,-0.07) | 0.14 (0.03,0.25) |
| SBP at age 24 years (mmHg) ^b^ | 122.07 (118.68,125.45) | -0.12 (-0.93,0.68) |

^a^Mean trajectory is centred on the sex-specific median of age at peak height velocity for each sex (age ~11.7 for females and age ~13.6 for males).

^b^ Estimated using regression coefficients for the intercept and rates of change per year spent in each spline period.

Adjusted for birth weight, gestational age, maternal education, parity, maternal smoking during pregnancy, maternal age, maternal pre-pregnancy BMI, household social class, marital status, partner education, breastfeeding, BMI residuals.

SBP, Systolic Blood Pressure; CI, confidence interval; mmHg/y, millimetres of mercury per year.

**eTable 11 Adjusted mean trajectory and mean difference in trajectory of SBP per year later age at peak height velocity, from chronological age multilevel models**

|  | **Mean trajectory (95% CI) of SBP^a^** | **Mean difference (95% CI) in SBP per year later aPHV** |
| --- | --- | --- |
| **Females** |  |  |
| SBP at 3 years of age (mmHg) | 92.70 (87.26,98.15) | 0.87 (-0.34,2.09) |
| Change in SBP 3-7 years (mmHg/y) | 1.45 (-0.57,3.48) | -0.37 (-0.82,0.08) |
| Change in SBP 7-12 years (mmHg/y) | 1.46 (0.94,1.98) | -0.35 (-0.47,-0.24) |
| Change in SBP 12-16 years (mmHg/y) | 3.63 (2.79,4.46) | 0.56 (0.37,0.74) |
| Change in SBP 16-20 years (mmHg/y) | -3.47 (-5.21,-1.73) | -0.16 (-0.55,0.22) |
| Change in SBP 20-24 years (mmHg/y) | 1.11 (-0.11,2.33) | -0.04 (-0.31,0.23) |
| SBP at age 24 years (mmHg) | 110.90 (108.17,113.63) | -0.94 (-1.53,-0.34) |
| **Males** |  |  |
| SBP at 3 years of age (mmHg) | 87.31 (82.15,92.47) | -0.94 (-2.00,0.13) |
| Change in SBP 3-7 years (mmHg/y) | 3.10 (1.16,5.05) | 0.32 (-0.08,0.72) |
| Change in SBP 7-12 years (mmHg/y) | 1.49 (0.98,2.00) | -0.33 (-0.43,-0.23) |
| Change in SBP 12-16 years (mmHg/y) | 5.97 (5.13,6.80) | -0.14 (-0.30,0.02) |
| Change in SBP 16-20 years (mmHg/y) | -2.88 (-4.86,-0.90) | 0.81 (0.42,1.20) |
| Change in SBP 20-24 years (mmHg/y) | 1.01 (-0.44,2.46) | -0.28 (-0.57,0.01) |
| SBP at age 24 years (mmHg) | 121.96 (118.47,125.45) | -0.04 (-0.72,0.65) |

^a^Mean trajectory is centred on the sex-specific median of age at peak height velocity for each sex (age ~11.7 for females and age ~13.6 for males).

^b^ Estimated using regression coefficients for the intercept and rates of change per year spent in each spline period.

Adjusted for birth weight, gestational age, maternal education, parity, maternal smoking during pregnancy, maternal age, maternal pre-pregnancy BMI, household social class, marital status, partner education, breastfeeding, BMI residuals.

SBP, Systolic Blood Pressure; CI, confidence interval; mmHg/y, millimetres of mercury per year.
